# Propranolol reduces Parkinson’s tremor and inhibits tremor-related activity in the motor cortex: a placebo-controlled crossover trial

**DOI:** 10.1101/2024.08.01.24311354

**Authors:** Anouk van der Heide, Maaike Wessel, Danae Papadopetraki, Dirk E.M. Geurts, Teije H. van Prooije, Frank Gommans, Bastiaan R. Bloem, Michiel F. Dirkx, Rick C. Helmich

**Affiliations:** Department of Neurology, Centre of Expertise for Parkinson & Movement Disorders, Radboud University Medical Centre, Nijmegen, the Netherlands; Donders Institute for Brain, Cognition, and Behavior, Centre for Cognitive Neuroimaging, Radboud University Nijmegen, the Netherlands; Department of Psychiatry, Radboud University Medical Centre, Nijmegen, the Netherlands; Department of Cardiology, Maxima Medical Centre, Veldhoven, the Netherlands

**Keywords:** Parkinson’s disease, tremor, propranolol, noradrenergic system, neuroimaging

## Abstract

**Objective:** Parkinson’s disease (PD) resting tremor is thought to be initiated in the basal ganglia and amplified in the cerebello-thalamo-cortical circuit. Since stress worsens tremor, the noradrenergic system may play a role in amplifying tremor. We tested if and how propranolol, a non-selective beta-adrenergic receptor antagonist, reduces PD tremor, and whether or not this effect is specific to stressful conditions.

**Methods:** In a cross-over, double-blind intervention study, participants with PD resting tremor received propranolol (40mg, single dose) or placebo (counter-balanced) on two different days. During both days, we assessed tremor severity (with accelerometry) and tremor-related brain activity (with functional Magnetic Resonance Imaging; fMRI), as well as heart rate and pupil diameter, while subjects performed a stressful cognitive load task that has been linked to the noradrenergic system. We tested for effects of drug (propranolol vs. placebo) and stress (cognitive load vs. rest) on tremor power and tremor-related brain activity.

**Results:** We included 27 PD patients with clear resting tremor. Tremor power significantly increased during cognitive load vs. rest (*F*(1,19)=13.8; *p*=.001; η_p_^2^=0.42) and decreased by propranolol vs. placebo (*F*(1,19)=6.4; *p*=.02; η_p_^2^=0.25), but there was no interaction. We observed task-related brain activity in a stress-sensitive cognitive control network, and tremor power-related activity in the cerebello-thalamo-cortical circuit. Propranolol significantly reduced tremor-related activity in the motor cortex compared to placebo (*F*(1,21)=5.3; *p*=.03; η_p_^2^=0.20), irrespective of cognitive load.

**Interpretation:** Our findings indicate that the noradrenergic system has a general, context-independent role in amplifying PD tremor at the level of the primary motor cortex.

## Introduction

Parkinson’s disease (PD) is characterized by progressive degeneration of the main ascending neurotransmitters systems, including the dopaminergic, noradrenergic, and serotonergic systems^1, 2^. While nigro-striatal dopamine depletion is related to bradykinesia and rigidity, this association is less clear for resting tremor^3–5^. Clinical evidence suggests an additional role for the noradrenergic system in the pathophysiology of PD tremor. Specifically, PD tremor increases during experimentally induced stress, such as cognitively demanding tasks^6, 7^, and patients rate tremor as their most stress-sensitive symptom^8^. Furthermore, while levodopa is effective in reducing tremor^9–11^, this effect is diminished by stressful contexts^12^. This highlights an important clinical problem: people experience most tremor during psychological stress, when dopaminergic treatment is least effective. These observations suggest that the noradrenergic system is involved in the pathophysiology of PD tremor, but the mechanisms are unclear.

According to the ‘dimmer-switch hypothesis’ of PD tremor, pathological activity in the striato-pallidal circuit triggers tremor onset analogous to a *light switch*^13^, while increased activity in the cerebello-thalamo-cortical circuit amplifies tremor power analogous to a *light dimmer*. We have previously shown that cognitive load, which activates the noradrenergic system, increases PD tremor through excitatory influences on the thalamus^6^. Other findings suggest that the involvement of the noradrenergic system in PD tremor may not be restricted to stressful situations: spontaneous fluctuations in tremor power in the absence of any task were tightly correlated with fluctuations in heart rate and pupil diameter, i.e. two proxy measures of noradrenergic activity^14^. Indeed, imaging and post-mortem studies have shown that the integrity of the locus coeruleus (LC), the main cerebral source of noradrenaline, is associated with the presence or absence of PD tremor^15, 16^. While noradrenergic terminal function was decreased in PD patients versus controls, noradrenergic projections from the LC towards the thalamus were more preserved in patients with clear tremor^15^. Accordingly, lesioning noradrenergic LC terminals in a PD rat model reduced the development of tremor^17^. In non-PD tremor disorders, such as essential tremor, drugs acting on the noradrenergic system influence tremor: sympathomimetics increase tremor^18^, while beta-blockers reduce tremor^19^. Some studies have suggested that beta-blockers may improve PD tremor as well^20–24^, but not all evidence points in the same direction^25, 26^ (**Supplementary table 1**). These studies most frequently administered propranolol, a non-selective beta-adrenergic receptor antagonist. Propranolol decreases the activity of the sympathetic nervous system, thus reducing the release of noradrenaline in the brain^27^.

Here, we used pharmacological imaging to parse the role of the noradrenergic system in the pathophysiology of PD tremor, and to test if this role is specific to stressful conditions or not. We performed a placebo-controlled, cross-over trial to investigate the effect of propranolol on tremor severity (accelerometry) and tremor-related activity (concurrent accelerometry-fMRI). Based on previous findings^6^, we hypothesized that propranolol would reduce resting tremor particularly during cognitive load, by inhibiting tremor-related activity in the thalamus. We also tested the alternative hypothesis that the effects of propranolol on tremor generalize across stressful and resting conditions, given evidence for the role of noradrenergic mechanisms in PD tremor at rest^14^.

## Methods

### Study population

We included 34 people with PD according to the Movement Disorders Society (MDS) criteria^28^, with a clear resting tremor in at least one arm (MDS Unified PD Rating Scale [MDS-UPDRS] III tremor-score ≥2; range 0-4). Exclusion criteria were: neurological or current psychiatric comorbidities; contraindications for MRI; cardiac arrhythmias; possible intolerance for beta-blockers (bradycardia, peripheral vascular diseases, diabetes mellitus, chronic obstructive pulmonary disease, asthma, hypotension); use of medication that may interact with propranolol or influences its metabolism; severe head tremor or dyskinesias; or cognitive impairment (Mini-Mental State Examination [MMSE] score <26)^29^.

Participants underwent an extensive phone screening prior to inclusion. Given the exploratory nature of this study and the lack of similar approaches, we did not perform a formal power calculation. Our desired sample size (*N*=24 with MRI) was based on previous studies reporting the effects of (dopaminergic) pharmacological interventions on tremor-related brain activity^10, 30^.

Measurements took place at the Donders Center for Cognitive Neuroimaging in Nijmegen (March 2020-September 2021). We performed electrophysiological tremor assessments in 27 participants, of whom 24 underwent successful fMRI scanning during both sessions (one patient excluded from analyses; final sample *N*=23, see **Figure 1**). **Table 1** shows the characteristics of participants included in data analysis (*N*=27).

**Figure 1.**
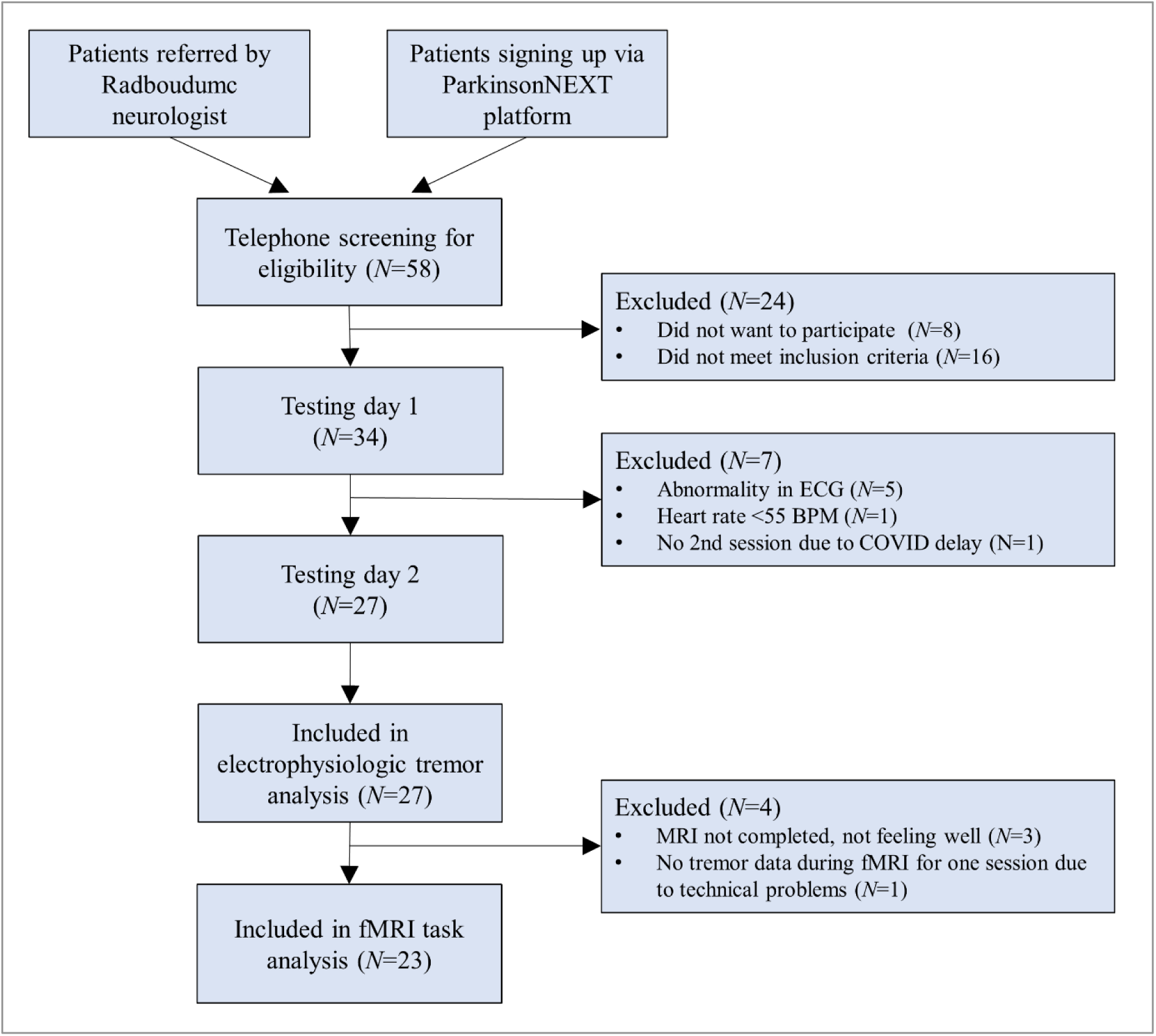
Participant flow chart. This participant flow chart shows how many participants were screened from the two recruitment sources, how many were eligible and how many were included per testing day and per analysis.

**Table 1.** Clinical characteristics.

| Characteristic | Mean (±SD) / N |
| --- | --- |
| Sex (Female \| Male) | 9 \| 18 |
| Age (years) | 61.4 (8.2) |
| Disease duration (years) | 4.2 (2.9) |
| Most affected side (Right \| Left) | 17 \| 10 |
| Dopaminergic medication user (Yes \| No) | 26 \| 1 |
| LEDD (mg/day) | 598.1 (386) |
| Mini Mental State Examination (total score) | 28.8 (1.3) |
| Hoehn and Yahr stage (HY 1 \| HY 2 \| HY 2.5) | 4 \| 22 \| 1 |
| MDS-UPDRS III (total score, OFF medication) | 28.8 (10.8) |
| Tremor total | 9.3 (4.4) |
| Resting tremor | 6.1 (2.6) |
| Postural tremor | 1.9 (1.4) |
| Kinetic tremor | 1.2 (1.1) |
| Non-tremor total | 19.5 (8.2) |
| Bradykinesia & rigidity | 13.7 (6.9) |
| Axial symptoms | 5.9 (1.9) |

Disease characteristics of participant ( $N=27$ ) included in the analysis of the electrophysiological tremor assessment. Some of these ( $N=4$ ) were not included in the fMRI-analysis. LEDD = levodopa equivalent daily dosage; MDS-UPDRS = Movement Disorders Society Unified Parkinson's Disease Rating Scale. Tremor total = MDS-UPDRS III items 15-18 (Resting tremor = items 17+18; Postural tremor = item 15; Kinetic tremor = item 16); Non-tremor total = MDS-UPDRS III items 1-14 (Bradykinesia & rigidity = items 3 & 4-8; Axial symptoms = items 1-3 & 9-14).

### Ethical approval and patient recruitment

Participant recruitment was performed through the Radboud University Medical Centre Neurology department and through patient recruitment platform ParkinsonNEXT (https://www.parkinsonnext.nl). All participants gave written informed consent prior to the experiment. The local medical ethical committee (METC Arnhem-Nijmegen) approved all procedures and communication materials (reference 2016-3101; NL59724.091.16). The EudraCT number is 2016-004629-18, and the study was also registered in the ISRCTN registry (ISRCTN89589002).

### Experimental design

Our cross-over, double-blind, placebo-controlled design consisted of two testing days of ∼5 hours. Participants received propranolol (40 mg, dispersed in water) on one day, and a placebo (cellulose dispersed in water) on the other (order counterbalanced, **Supplementary notes**). Each testing day consisted of clinical and behavioral tests and fMRI (**Figure 2A**).

**Figure 2.**
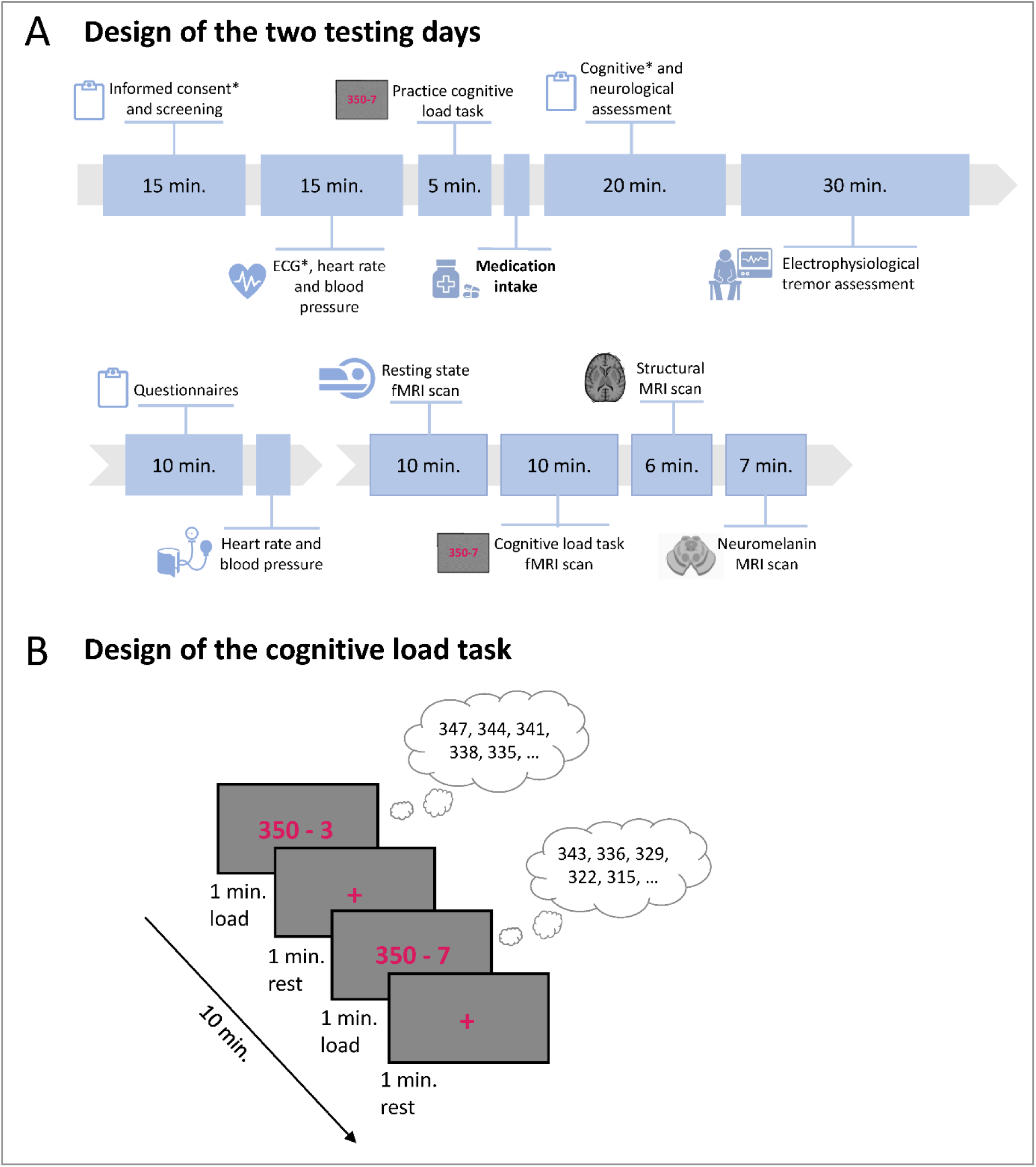
Study design and cognitive load task. **(A)** shows an overview of the testing day. Each session consisted of a behavioral part and a part in the MRI scanner. **(B)** visualizes the cognitive load task that was performed during fMRI scanning. *: only done during first testing day. fMRI = functional Magnetic Resonance Imaging.

At the start of the first visit, an electrocardiogram (ECG) was recorded and checked by a physician for irregularities in rhythm and conductance that could indicate contraindications for propranolol. Blood pressure and heart rate were monitored. Participants were in a practically defined OFF-state during all measurements (>12 hours after the last dose of levodopa, >24 hours after dopamine agonists).

### Measurements

#### General clinical measurements

During both sessions, motor symptoms were assessed using the MDS-UPDRS III^31^. Cognitive function was assessed during the first session with the MMSE^32^.

#### Electrophysiological tremor assessment and preprocessing

We performed an electrophysiological tremor assessment, with four conditions: rest (forearms on the armrests, hands unsupported), cognitive load (serial-subtraction task), postural tremor (arms stretched forward) and kinetic tremor (slow wrist extension and flexion of the most-affected arm). All conditions (each 1 minute) were repeated twice. Participants were told to never voluntarily suppress the tremor. MRI-compatible EMG-electrodes were placed on both forearm muscles (extensor digitorum communis and flexor carpi radialis), and, in case of leg tremor, on the tibialis anterior and gastrocnemius. An MRI-compatible tri-axial accelerometer (Brain Products; sampling frequency (Fs) = 5,000Hz) was placed on the dorsum of the most-affected hand.

Preprocessing involved detrending and demeaning the signal to remove temporal drifts and band-pass filtering of 1-20Hz to filter out noise, as done before^11^. We used Fieldtrip to calculate power spectra for each condition and each accelerometry channel (x/y/z) between 1-16Hz using a 5-second Hanning taper, resulting in a 0.2Hz spectral resolution. Only fluctuations within the frequency ranges of 3.5-7Hz for resting tremor and 3.5-10Hz for postural and kinetic tremor were considered as tremor-related activity. The log10-transformed peak power was determined for the channel with the highest power.

#### MRI image acquisition and preprocessing

fMRI was performed on a Siemens PRISMA 3T MRI-system with a 32-channel head-neck coil. We used a multi-band echo planar imaging sequence with multi-band acceleration factor 6, repetition time 1000ms, echo time 34ms, 72 axial slices, 2.0mm isotropic voxels and field of view 210mm (∼10 minutes, 600 images). Anatomical images were acquired using a magnetization-prepared rapid gradient-echo sequence, repetition time 2300ms, echo time 3.03ms, 192 sagittal slices, 1.0mm isotropic voxels and field of view 256mm (∼5 minutes). Participants were instructed to lie still with their eyes open (confirmed with online eye-tracking).

Preprocessing of functional and structural images was done using fMRIprep v20.2.1^33^. Preprocessing steps for structural scans involved intensity nonuniformity correction, skull-stripping, brain surface reconstruction, spatial normalization to Montreal Neurological Institute (MNI) space, and brain tissue segmentation. Preprocessing for the functional runs included correction for motion-related variance using ICA-AROMA, nuisance regression of average cerebrospinal fluid and white matter time series together with 24 motion derivatives, and high-pass filtering (>0.007Hz). ICA-AROMA components were visually checked by two researchers and manually corrected if necessary. The first five images of functional runs were discarded. Then functional scans were co-registered to the T1-weighted reference image and mapped to MNI space. Preprocessed functional images were spatially smoothed using a Gaussian kernel of 6mm full width at half-maximum using SPM12 (https://www.fil.ion.ucl.ac.uk/spm/software/spm12).

#### Functional MRI task: cognitive load task

Participants performed a validated 10-minute stress-inducing cognitive load task during both sessions (propranolol and placebo). This task consisted of alternating 1-minute blocks of mental arithmetic (cognitive load) and observation of a cross (rest; **Figure 2B**)^6^. Participants were instructed to perform the mental calculations (in silence) as fast as possible and to start again if they reached zero. We used continuous eye-tracking of the left eye (Eyelink 1000 plus, Fs = 1,000Hz) to monitor pupil diameter, and we measured heart rate using a pulse oximeter on the thumb of the less-affected hand (Blood Pulse Sensor; Brain Products), and we continuously measured tremor during the task as outlined below^6, 12^. After the task, participants rated their perceived stress level, separately for cognitive load and rest blocks, using a Visual Analog Scale (range 1-5).

#### Debriefing

After the final testing day, all participants were asked which medication they thought they had received that day.

### Statistical analysis

We tested for effects of DRUG (propranolol vs. placebo), BLOCK (cognitive load vs. rest), and their interaction, on (1) tremor power (accelerometry or EMG, both inside and outside the scanner), (2) stress measures (pupil diameter, heart rate and perceived stress), and (3) tremor-related brain activity (BOLD). We tested the hypothesis that propranolol reduces tremor-related activity and tremor power specifically during stress (BLOCK*DRUG interaction). Analyses were performed using R-4.2.1 (www.r-project.org).

#### (1) Tremor power

Resting tremor power was calculated at each individual’s tremor peak, log10-transformed, and averaged across repetitions. For postural and kinetic tremor, we performed separate one-sided paired t-tests comparing the effect of DRUG (propranolol vs. placebo) on tremor peak power. We did the same analyses with tremor frequency as dependent variable. For one participant, only tremor frequency was compared between sessions; tremor power could not be computed due to technical issues. Inside the fMRI scanner, accelerometry data were unavailable in three participants due to technical issues.

#### (2) Stress measures

We performed two-way rm-ANOVA’s to test the effects of BLOCK (rest vs. cognitive load) and DRUG (propranolol vs. placebo) on heart rate, pupil diameter and perceived stress level. Pupil size data during fMRI were of insufficient quality in one or both sessions for ten participants, due to dropping of the eye lid, insufficient light, or inability to visualize the pupil. Heart rate data were of insufficient quality in nine participants.

#### (3) Tremor-related brain activity (fMRI)

At the first (subject-specific) level, we performed a multiple regression analysis in SPM12 with separate regressors modeling rest and cognitive load conditions for each session. As done before, we modelled tremor-related activity by adding scan-by-scan tremor power (accelerometry) and its first temporal derivative as two parametric modulation regressors for each of these four conditions^6^. To remove non-neural and motion-related noise, we included covariates of no interest (calculated by fMRIprep^33^) for framewise displacement, standardized DVARS (derivative of root-mean-square variance over voxels), cerebrospinal fluid, white matter signal, 24 head-motion derivatives and ICA-AROMA components to the first-level contrasts. For each individual, this resulted in four contrast-images depicting tremor-related activity for each condition (rest vs. cognitive load; propranolol vs. placebo), and in separate contrast images depicting task-related activity (cognitive load>rest; propranolol vs. placebo). For participants with left as most-affected hand (*N*=9), we flipped the contrast images in the axial plane for tremor power-related and tremor change-related contrasts.

At the second (group) level, we first verified that individuals showed task-related activity in a cognitive control network (cognitive load>rest, averaged across medication sessions) and tremor-related activity in the cerebello-thalamo-cortical circuit (averaged across blocks and medication sessions), as done before^6^. We used Threshold Free Cluster Enhancement (TFCE) in SPM12 (http://dbm.neuro.uni-jena.de/tfce/) and assessed significance with non-parametric permutation testing (10,000 permutations). We corrected for multiple comparisons (threshold p<.05), familywise error (FWE) corrected at the voxel-level for all analyses. Corrections were done across the whole brain for task-related activity, and within regions of interest (ROI; **Table 2**) in the cerebello-thalamo-cortical circuit for tremor-related activity, given previous work^13^. Mean framewise displacement for each participant and session was added as covariate. We used the SPM Anatomy Toolbox 2.1 for anatomical localization of activity clusters^34^.

**Table 2.**
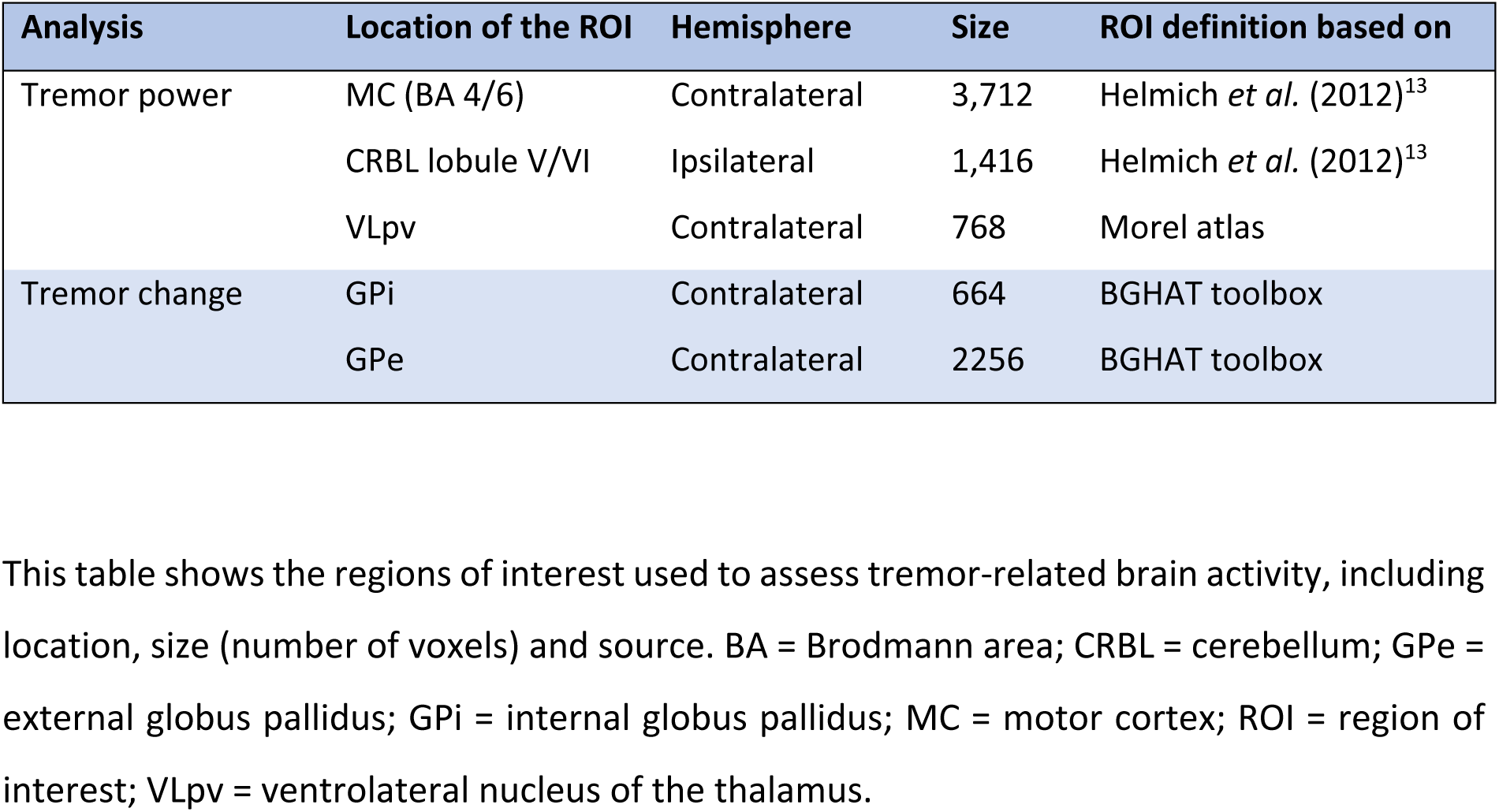
Regions of interest used to assess tremor-related brain activity.

Second, we used rm-ANOVA’s to test for effects of BLOCK (rest vs. cognitive load) and DRUG (propranolol vs. placebo) on tremor-related activity within the cerebello-thalamo-cortical circuit (average beta-values depicting tremor-related activity in each ROI). We also performed an exploratory whole brain search.

Finally, we assessed the relationship between drug-related changes in tremor power, and drug-related changes in tremor-related brain activity with Spearman correlations (R-4.2.1). For this, we calculated the *change* in mean beta-values for each ROI showing effects of propranolol, and the *change* in tremor power, per participant.

We also report Bayesian statistics to test the validity of null responses, particularly regarding our main hypothesis (BLOCK*DRUG interaction on tremor-related brain activity and resting tremor power). Bayes factors (BF_10_) of 1–3, 3–10 or >10 were respectively considered anecdotal, moderate or strong evidence for the alternative hypothesis, whereas BF_10_ of 0.33– 1, 0.1–0.33 or <0.1 were considered anecdotal, moderate or strong evidence for the null hypothesis (**Supplementary notes**).

## Results

### Tremor power

Outside the scanner, cognitive load *increased* rest tremor power (main effect BLOCK: F(1,24)=10.5; p=.003; η_p_^2^=0.31), whereas propranolol *reduced* rest tremor power (main effect DRUG: F(1,24)=10.6; p=.003; η_p_^2^=0.31) (**Figure 3A**). We found no BLOCK*DRUG interaction (F(1,24)=0.0; p=.95; η_p_^2^=0.00; BF_10_=0.27). Rest tremor frequency was unaffected by propranolol (main effect DRUG: F(1,25)=0.7; p=.42; η_p_^2^=0.03), or cognitive load (main effect BLOCK: F(1,25)=3.0; p=.10; η_p_^2^=0.11); no interaction (BLOCK*DRUG: F(1,25)=0.1; p=.74; η_p_^2^=0.00). Propranolol also reduced postural tremor power (t(16)=3.6; p=.003; Cohen’s d=0.86) (**Figure 3B**), but not its frequency (t(17)=1.1; p=.28; Cohen’s d=-0.27). Kinetic tremor was present across both sessions in only ten participants. Propranolol did not affect tremor power (t(9)=0.1; p=.89; Cohen’s d=0.05) (**Figure 3C**) or frequency (t(9)=0.7; p=.51; Cohen’s d=0.22).

**Figure 3.**
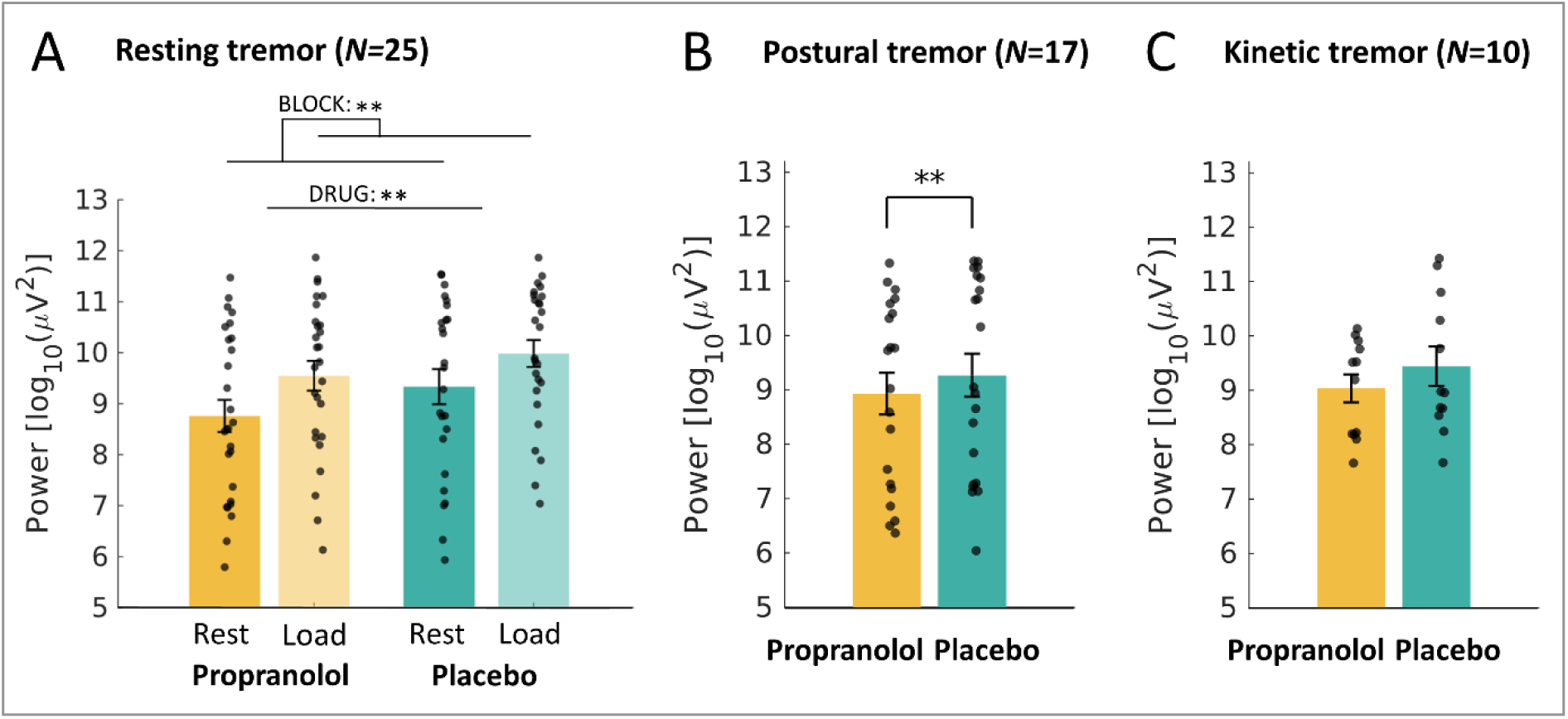
Effect of propranolol on different types of PD tremor. The effect of propranolol on average tremor power (± SEM), measured with accelerometry during two one-minute blocks per condition, for **(A)** resting tremor, **(B)** postural tremor and **(C)** kinetic (action) tremor. *: p<.05, **: p<.01, ***: p<.001.

During MRI, we observed the same effects as outlined above. Cognitive load *increased* rest tremor power (main effect BLOCK: F(1,19)=13.8; p=.001; η_p_^2^=0.42), while propranolol *reduced* tremor power (main effect DRUG: F(1,19)=6.4; p=.02; η_p_^2^=0.25; **Figure 4A**). There was no BLOCK*DRUG interaction (F(1,19)=0.7; p=.41; η_p_^2^=0.04; BF_10_=0.50).

**Figure 4.**
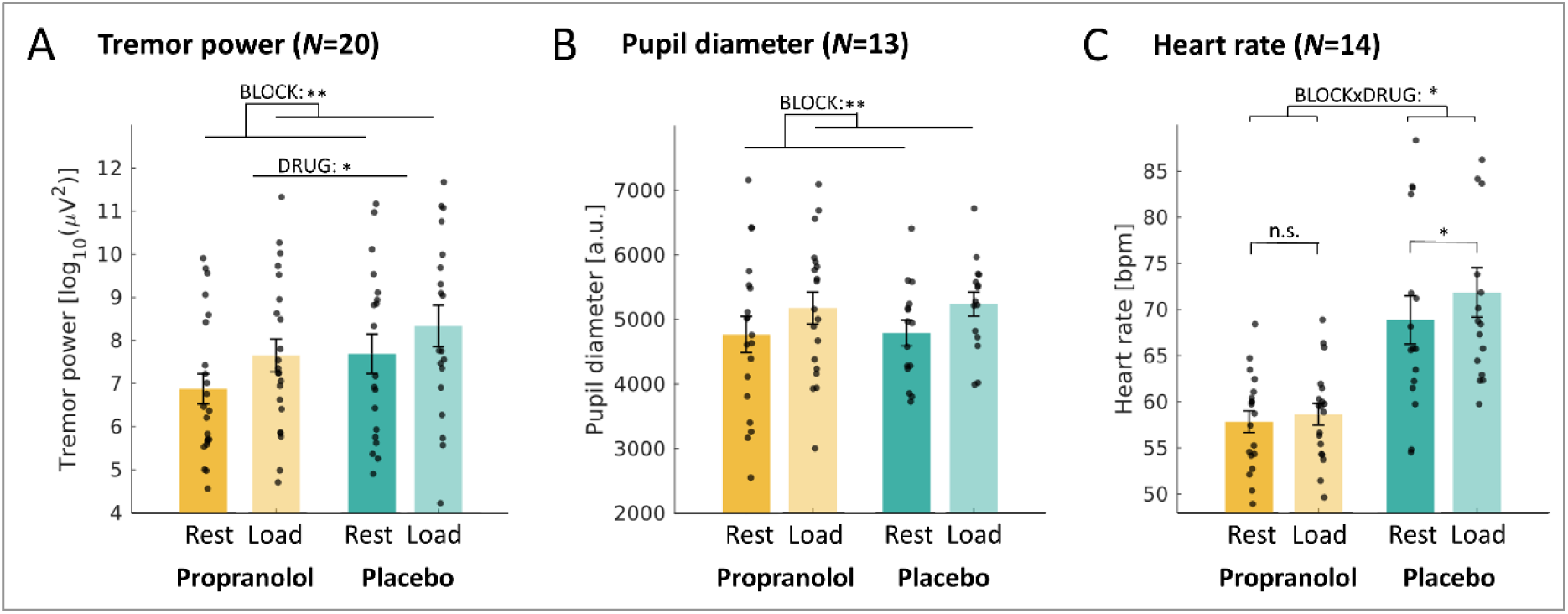
Effects of propranolol and cognitive load on physiological measures. The effect of cognitive load and propranolol intake on **(A)** tremor power, **(B)** pupil diameter and **(C)** heart rate during the fMRI cognitive load task. *: p<.05, **: p<.01, ***: p<.001.

### Stress measures

Inside the scanner, pupil size increased during cognitive load (main effect BLOCK: F(1,12)=19.2; p=.001; η_p_^2^=0.62), but there was no effect of propranolol (main effect DRUG: F(1,12)=0.1; p=.78; η_p_^2^=0.01) (**Figure 4B**) and no interaction (F(1,12)=0.0; p=.85; η_p_^2^=0.00; BF_10_=0.41). For heart rate, there was a significant BLOCK*DRUG interaction (F(1,13)=5.3; p=.04; η_p_^2^=0.29). Post-hoc tests showed that heart rate increased for cognitive load compared to rest during the placebo session (*t*(13)=2.8; *p*=.02; Cohen’s d=0.75), but not during the propranolol session (*t*(13)=1.1; *p*=.29; Cohen’s d=0.30) (**Figure 4C**). Participants perceived the cognitive load blocks as more stressful than rest blocks (average score 2.7 vs. 2.0 [range 1-5]; main effect BLOCK: *F*(1,22)=19.4; *p*=.000; η_p_^2^=0.47). They perceived more stress during the placebo session (average score 2.6 vs. 2.0; main effect DRUG: *F*(1,22)=6.7; *p*=.02; η_p_^2^=0.23). There was no interaction (*F*(1,22)=0.6; *p*=.44; η_p_^2^=0.03).

### Effects of cognitive load on brain activity

Cognitive load was associated with *increased* activity in a cognitive control network, including the posterior-medial frontal cortex, middle frontal cortex, superior and inferior parietal lobule, superior temporal lobe, and cerebellum (**Figure 5A**), anatomical details in **Supplementary table 2**. Propranolol had no effect on task-related activity, and there was no BLOCK*DRUG interaction.

**Figure 5.**
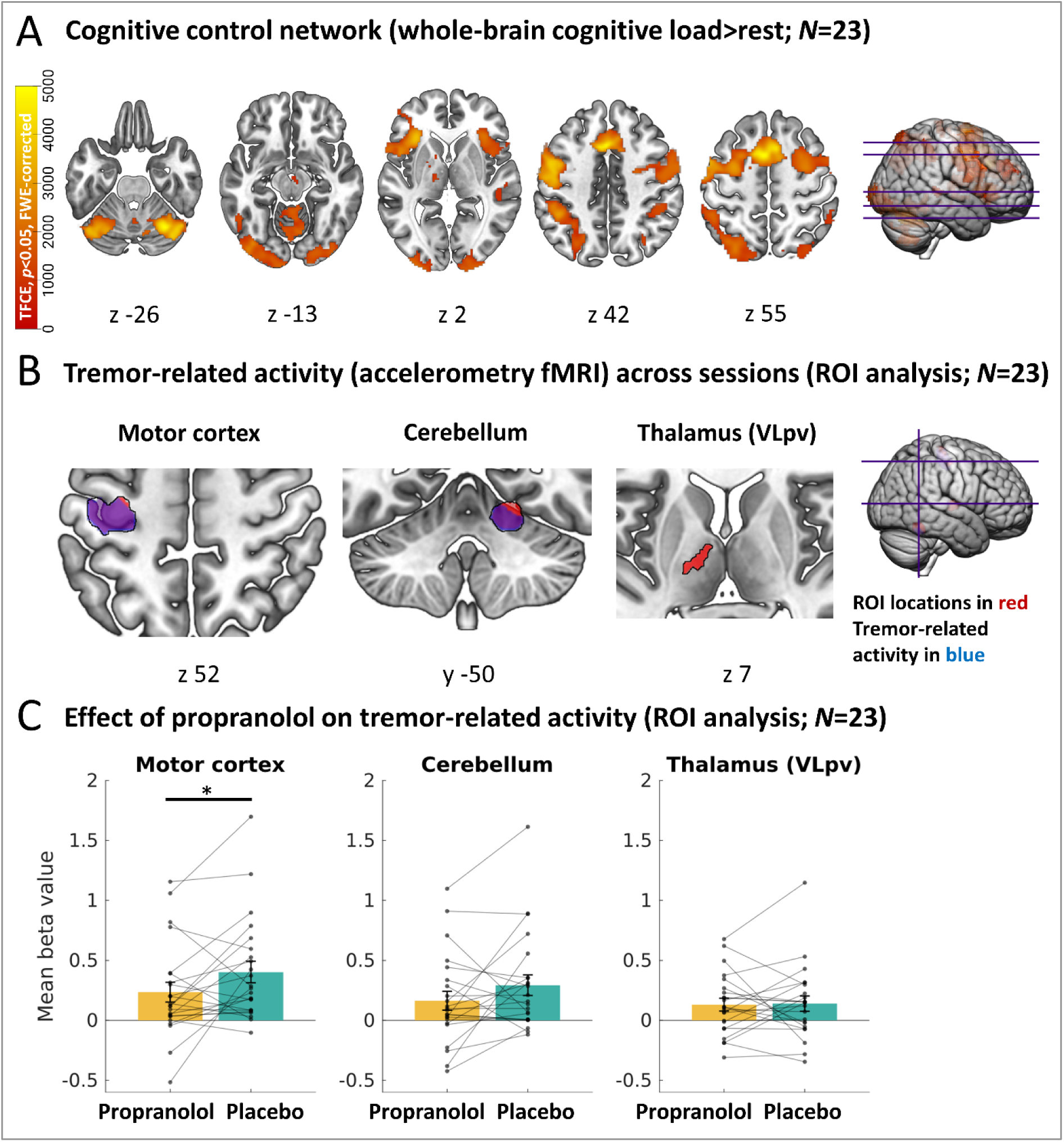
Cerebral activity patterns measured by functional MRI. (A) shows the activation of a cognitive control network during the cognitive load task (whole-brain cognitive load>rest, averaged across sessions). The image shows TFCE-values of significant clusters, FWE-corrected. (B) shows tremor power-related activity across blocks in the cerebello-thalamo-cortical network (ROI-based analysis; averaged across sessions and blocks). The image shows significant clusters, FWE-corrected. (C) shows the comparison of tremor-related activity during the placebo and propranolol session, averaged over significant voxels in the cerebello-thalamo-cortical network (ROI-based analysis; averaged across blocks). Bars represent mean beta values (±SEM) and dots show individual averaged beta values. *: p<.05, **: p<.01, ***: p<.001.

### Tremor-related brain activity

Across sessions and conditions, we replicated previous findings showing tremor power-related activity within the contralateral motor cortex, ipsilateral cerebellum and contralateral VLpv (the latter trend-significant) (**Figure 5B**). Anatomically, these clusters largely overlapped with clusters found in previous studies^6, 13^. We did not observe tremor change-related activity in the basal ganglia, similar to a previous study using the same task^6^ (**Supplementary table 2**). Propranolol significantly reduced tremor-related activity in the motor cortex independent of cognitive load (main effect DRUG: *F*(1,21)=5.3; *p*=.03; η_p_^2^=0.20; **Figure 5C**). In the cerebellum, the effect of propranolol approached significance (main effect DRUG: *F*(1,21)=3.1; *p*=.09; η_p_^2^=0.13). Propranolol did not influence tremor-related activity in the thalamus (VLpv; main effect DRUG: *F*(1,21)=0.0; *p*=.93; η_p_^2^=0.00). There were no main effects of BLOCK on tremor-related activity, and BLOCK*DRUG interactions were not significant (motor cortex: *F*(1,22)=1.3; *p*=.26; η_p_^2^=0.06; BF_10_=0.50; cerebellum: *F*(1,22)=0.2; *p*=.67; η_p_^2^=0.01; BF_10_=0.26; VLpv: *F*(1,22)=0.3; *p*=.57; η_p_^2^=0.01; BF_10_=0.38). Whole-brain analyses did not reveal tremor-related BLOCK or DRUG effects in regions outside the cerebello-thalamo-cortical circuit.

There was no correlation between session-specific changes (placebo>propranolol) in tremor power (accelerometry) and tremor-related brain activity in the motor cortex (fMRI): rho=0.12; p=0.63.

### Debriefing

After completion of the second testing day, 19 of the 27 participants guessed correctly in which order they had received the medication.

## Discussion

We investigated the role of the noradrenergic system in the pathophysiology of PD tremor, by testing if a behavioral *induction* of noradrenergic activity (cognitive load task) and a pharmacological *inhibition* of noradrenergic activity (propranolol) influenced both tremor power (accelerometry) and tremor-related brain activity (fMRI) in 27 people with PD resting tremor. Based on previous findings^6^, we hypothesized that propranolol would inhibit the stress-related increase in tremor power and tremor-related activity. There are two main findings. First, cognitive load increased tremor power, while propranolol reduced tremor power, independently of the task (no interaction). This suggests that PD tremor is modulated by noradrenergic activity: not only during stressful conditions, but also during “rest”. Second, propranolol reduced tremor amplitude-related activity in the contralateral motor cortex, but not in the ipsilateral cerebellum or the VLpv.

### Effects of propranolol on tremor power

Propranolol reduced tremor power across two separate measurements in different contexts, i.e. outside the scanner (seated) and inside the scanner (lying supine), suggesting this finding is robust. Our findings also suggest that the effect of propranolol on PD tremor is relevant: the magnitude of tremor *reduction* after propranolol was similar to the *increase* observed from rest to cognitive load blocks (η_p_^2^ effect-size of 0.3 for both effects). This worsening of tremor during cognitive load is a common and clearly visible observation in clinical practice, experienced as impactful by patients^35^.

The tremor-reducing effect of propranolol was not specific to the stressful (cognitive load) condition, but was also effective during rest periods. These findings are in line with a previous study in seven PD patients, where propranolol counteracted the tremor-increase caused by adrenaline injection, without impact on tremor-increase observed during mental calculations^36^. Effects of propranolol on tremor at rest were not assessed in that work. Our findings cannot be attributed to the cognitive load task failing to activate the noradrenergic stress system, as we observed clear task effects on proxies of (nor)adrenergic activity, i.e. increased heart rate (in the placebo session) and pupil diameter (**Figure 4**). Additionally, we observed task-related activity in a cognitive control network previously linked to psychological stress (**Figure 5A**)^37^. It is also unlikely that our study was underpowered to detect a BLOCK*DRUG interaction: Bayesian statistics convincingly supported the null hypothesis (BF<0.3). Instead, our findings suggest a more general role of the noradrenergic system in PD tremor, beyond stressful situations. This fits with recent findings showing that spontaneous tremor power fluctuations at rest correlate with fluctuations in proxies for noradrenergic activity (pupil and heart rate)^14^. The clinical implication of our findings is that propranolol could be a useful treatment option for PD tremor, whether or not it is triggered by stress.

Propranolol effectively reduced both resting tremor and postural tremor, but not kinetic tremor. This suggests that the noradrenergic system plays no role in kinetic tremor, although the small sample number of patients with kinetic tremor (*N*=10) warrants caution when interpreting this result.

### Propranolol inhibits tremor-related brain activity

Tremor was associated with a cerebral circuit comprising the contralateral motor cortex, ipsilateral cerebellum, and contralateral VLpv (exhibiting a trend toward significance). Propranolol significantly reduced tremor-related activity in the motor cortex, but not in the cerebellum (which was trend-significant), or the VLpv. Similar to the effects we observed on tremor power, but in contrast to our hypothesis, there was no BLOCK*DRUG interaction in any of these three regions. Bayesian evidence in favour of the null hypothesis was anecdotal to moderate for all regions (BF_10_=0.38-0.50), and with very small effect sizes (0.0-0.1). Hence, it is unlikely that studies in larger samples would detect a clinically relevant interaction effect.

Our finding that propranolol reduced tremor-related activity in the motor cortex, independent of the cognitive task, fits with the notion that the LC has anatomical projections to all nodes of the cerebello-thalamo-cortical circuit^38^. Furthermore, the motor cortex, with a particularly high glucocorticoid receptor density, is susceptible to the effects of stress^39^. In animal studies, application of noradrenergic antagonists over the primary motor cortex hyperpolarized layer-V output neurons and decreased firing rates^40^, which is in line with our findings.

Importantly, stress-related noradrenergic activity activates several receptors (including beta-1, beta-2, beta-3, and alpha-receptors), whereas propranolol only blocks beta-1 and beta-2 receptors^27^. The latter are differently expressed within the cerebello-thalamo-cortical circuit (**Supplementary figure**). Assessing the genetic expression of these receptors, derived from postmortem samples (Allen Brain Institute^41^; MNI-spatial maps at https://neurosynth.org/genes), showed a high genetic expression of beta-1 receptors in the motor cortex ROI (z=0.32) that is not seen in the VLpv (z=-0.41) or cerebellum (z=-0.08). On the other hand, the genetic expression of beta-2-receptors is found mainly in the VLpv (z=0.70), but not in the motor cortex (z=-0.22) or cerebellum (z=-0.45). Our finding that propranolol reduced tremor-related activity only in the motor cortex, therefore suggests that beta-1 receptors were primarily targeted, but further research should confirm this.

### Peripheral contributions

We cannot rule out that some of the clinical effects are explained by peripheral effects of propranolol. Although PD tremor is primarily generated by cerebral oscillations, it is well-accepted that peripheral mechanisms play an additional role. For example, PD tremor can be influenced by somatosensory afferents, since changes in tremor power may occur even after slight adaptations in limb posture^42^. However, studies showed that mechanical perturbations could only reset PD tremor in a subsample of participants^43^, or only in specific conditions^44^. Somatosensory afferents might also have a role in *stabilizing* the tremor rhythm, as was substantiated by a study showing that deafferentation of a tremulous arm did not affect tremor power or frequency^45^. Since propranolol has both peripheral and central effects, it may reduce tremor through a combined action on both^46^. Since the extent of peripheral contributions to PD tremor may vary between people, the effect of propranolol on tremor via the cerebello-thalamo-cortical circuit might differ as well. This may explain why we did not find a correlation between propranolol-induced tremor power reduction and the effect of propranolol on the motor cortex.

### Limitations

The main strength of our study is that our multi-modal design allowed us to address both the clinical effect of propranolol on tremor and the underlying mechanisms. Some limitations should be considered. First, our sample is modest, and drop-out was relatively high (32%), mostly due to stringent ECG-based exclusion criteria. Second, although the study was double-blind, 19 of the 27 participants guessed correctly in which order they had received the medication. Unblinding may have occurred due to people noticing the effects of propranolol on heart rate (which decreased by 12.4bpm) or tremor itself. Third, although our results suggest that propranolol did not affect tremor-related VLpv activity, Bayesian analyses could not rule out this effect completely, possibly because our fMRI multi-band scanning protocol is less sensitive to activity in subcortical regions, including the thalamus^47^.

### Clinical implications

This placebo-controlled study shows that propranolol is effective in reducing PD tremor across both stressful and resting conditions. Insights from this study may improve treatments that attenuate the influence of the noradrenergic system on PD tremor, based on behavioral (e.g. mindfulness-based interventions^8, 48^) or pharmacological suppression (e.g. propranolol) of the noradrenergic system. Our findings also suggest that this effect is mediated by beta-1 receptors in the motor cortex, which opens opportunities for more targeted noradrenergic interventions with fewer side effects than propranolol. Furthermore, since noradrenaline has been shown to exert long-term effects on the motor cortex by promoting long-term plasticity^49^, daily use of propranolol use may have beneficial effects that extend beyond the single-dose findings we report.

## Acknowledgements

We thank all our study participants for their time and commitment to the study, and Eva Klimars for her help in setting up the study. Rick Helmich was financially supported by a VENI (#91617077) and VIDI (#09150172010044) grant from the Netherlands Organization for Scientific Research (Netherlands).

## Author contributions

AH coordinated the clinical study, collected the data, performed analyses, interpreted the data and drafted the manuscript. MW was involved in data collection, performed analyses and revised the manuscript critically for intellectual content. DP assisted in study coordination, was involved in data collection and revised the manuscript critically for intellectual content. DG, TP and FG assessed ECGs during testing days to screen inclusion criteria, and revised the manuscript critically for intellectual content. BB contributed in designing the study and has revised the manuscript critically for intellectual content. MD contributed to interpretation of the data and has revised the manuscript critically for intellectual content. RH conceptualized and designed the study, assessed ECGs during testing days, contributed to interpretation of the data and was involved in drafting the manuscript. All have read and approved the final version of the manuscript.

## Data availability

All derived and anonymized individual data supporting these findings are openly available at the Donders Repository at https://data.ru.nl/collections/di/dccn/DSC_3024005.02_550. All scripts used to generate the analyses presented in this paper are publicly archived at https://github.com/AnoukvanderHeide/Propranolol_PD_tremor.

## Supplementary materials

Supplementary material is available online.

