## Supplementary material for "Propranolol reduces Parkinson’s tremor and inhibits tremor-related activity in the motor cortex: a placebo-controlled crossover trial"

**Supplementary table 1. Overview of studies on the effect of beta-blockers on PD tremor**

| Study | Type of beta-blockers | Sample size | Design | Effect |
| --- | --- | --- | --- | --- |
| Abramsky *et al.* 1971^1^ | Propranolol (60-100 mg for 8 weeks) + levodopa (2-4 gr) | *N*=36 | Control-group with only levodopa | Tremor was reduced compared to only levodopa: tremor decreased in 72% of participants for propranolol and in 33% for only levodopa. |
| Dowzenko *et al.* 1976^2^ | Oxprenolol (60-120 mg daily for 3 weeks) | *N*=12 | Placebo-controlled cross-over, order fixed | Tremor intensity reduced in 7/12 participants. |
| Foster *et al.* 1984^3^ | Nadolol (80-320 mg) | *N*=8 | Placebo-controlled | Tremor (rest, postural and intention) reduction compared to placebo, most effect for severe tremor. |
| Henderson *et al.* 1994^4^ | Propranolol (20, 40 and 80 mg) | *N*=11 | Placebo-controlled | Tremor amplitude reduced by >30% in 5/11 participants, but did not outperform placebo and levodopa. |
| Herring 1964^5^ | Pronethalol (50 mg) | *N*=10 | Placebo-controlled (injection of saline) | Electrical muscle activity reduced to 31% of baseline, compared to 66% of baseline for water. |
| Kissel 1974^6^ | Propranolol + levodopa (>60 mg daily for 6-24 months) | *N*=25 | Cross-over, not blinded (also studying practolol, pindolol and quinidine) | Tremor reduced in 23/25 participants after combined therapy, better effect than either of the drugs alone. Practolol had no effect. |
| Koller & Herbster 1987^7^ | Propranolol (160 mg) | *N*=10 | Double-blind cross-over (also studying primidone & clonazepam) | Tremor amplitude reduced by 70% for resting tremor and by 50% for postural tremor. |
| Marsden & Owen 1967^8^ | Propranolol (80 mg) and pronethalol (200 mg) | *N*=8 | Not controlled, not blinded | Propranolol reduced adrenaline-induced tremor escalation but did not impact tremor-increase due to mental stress. |
| Owen & Marsden 1965^9^ | Propranolol (30 mg daily for a week) and pronethalol | *N*=16 | Placebo-controlled | Tremor was reduced compared to placebo, but degree of reduction varied considerably. |
| Strang 1965^10^ | Propranolol (up to 60 mg daily for 1-8 weeks; additionally, 20-50 mg intravenous in *N*=6) | *N*=26 | Not controlled, not blinded | Tremor was reduced by 10-20% in only three participants. Intravenous treatment did not affect tremor amplitude or rate. |
| Vas 1966^11^ | Propranolol (10 mg; intravenous) | *N*=10 | Double-blind placebo-controlled | Tremor amplitude was not significantly reduced. |

This table provides a summary of research studies with human participants, investigating the effects of beta-blockers to treat PD tremor. The data were retrieved in June 2023, and only trials are included for which an English abstract (or full article) was available. Studies included in this table administered either only a beta-blocker or a beta-blocker in combination with other parkinsonian medication.

**Supplementary note 1. Information regarding random allocation sequence**

To ensure unbiased allocation, the following approach was used. Two independent and qualified external researchers, unaffiliated with the study, were entrusted with the task of creating randomization lists. They ran a script that created the randomization list for assigning the medication for each specific session and subject combination. The same script was used to inform which medication should be prepared on each testing day.

The script was content-obscured, executable-only and solely accessible to the researchers assigned with preparing the medication. Crucially, the script also thoroughly documented every instance it was executed and by whom. This method ensured double-blindness and made it possible that the medication was prepared prior to every session. Notably, no blocking or block size was incorporated into the randomization process, maintaining simplicity.

These same external researchers were also responsible for preparing the study medication, which was securely stored in a locked cupboard. Propranolol, the active drug, was individually packaged for each participant by the pharmacy and arranged sequentially (participant numbers printed on each package). The placebo medication (cellulose) consisted of a single large box to accommodate all participants. The primary researcher collaborated closely with the external researchers, providing them with the corresponding subject numbers.

**Supplementary note 2. Bayesian statistics**

In addition to classical statistics, we reported Bayesian statistics using JASP 0.16.3^12^ for Bayesian analysis. Specifically, for reported non-significant effects, we tested for evidence in favor of the null hypothesis. Bayes factors quantify the ratio of accumulated evidence for the null hypothesis and the alternative hypothesis. We report Bayes factors in favor of the alternative hypothesis over the null hypothesis (BF_10_).

The reported Bayes factors were interpreted according to the JASP guidelines^12^: Bayes factors (BF_10_) of 1–3, 3–10 or >10 were respectively considered anecdotal, moderate or strong evidence for the alternative hypothesis, whereas BF_10_ of 0.33–1, 0.1–0.33 or <0.1 were considered anecdotal, moderate or strong evidence for the null hypothesis.

In order to evaluate the evidence against the interaction effects in the performed Bayesian repeated measures ANOVAs, we divided the BF_10_ of the model with the interaction by the BF_10_ of the model with only the two main effects (i.e. everything except the interaction).

By reporting these Bayes factors, we can substantiate statistical evidence in favor of *no effect* between groups, suggesting that the lack of a statistical difference in our conventional analysis was not driven by a lack of statistical power.

**Supplementary table 2. Brain activity related to cognitive load and tremor**

| **Cognitive control network (cognitive load>rest, averaged over sessions; whole brain analysis)** | | | | | |
| --- | --- | --- | --- | --- | --- |
| **Anatomical label (area)** | **Hemisphere** | **Cluster size (voxels)** | **TFCE value** | **MNI coordinates peak voxel (x, y, z)** | ***p* (FWE)** |
| Posterior-Medial Frontal | Medial | 19,224 | 5686.3  5063.8  5042.2 | -3, 9, 50  -1, 15, 44  -1, 5, 64 | <.001  <.001  <.001 |
| Cerebellum (Lobule VI and VIIa crus I) | Right | 9,286 | 4998.3  4181.0  3932.3 | 31, -63, -28  37, -69, -28  35, -51, -28 | <.001  <.001  <.001 |
| Inferior Parietal Lobule (hIP1) | Left | 3,346 | 3135.3  3087.3  2485.8 | -42, -43, 40  -46, -43, 48  -25, -67, 50 | <.001  <.001  <.001 |
| Superior Parietal Lobule (7A) | Right | 743 | 1483.6  1373.0  1360.6 | 17 -69 62  31, -69, 62  31, -65, 44 | .003  .005  .005 |
| SupraMarginal Gyrus ( hIP2) | Right | 563 | 1548.9  1466.6  1457.9 | 47, -37, 44  41, -43, 46  43, -33, 38 | .003  .004  .004 |
| Middle Frontal Gyrus | Right  Left | 829  3 | 1088.7  1023.0  1005.7  766.1 | 35, 38, 32  35, 40, 18  45, 42, 18  -31, 34, 28 | .012  .016  .017  .039 |
| Inferior frontal Gyrus | Left | 11 | 715.6 | -31, 27, -16 | .048 |
| Superior Temporal Gyrus | Right | 207 | 909.4  899.5  896.9 | 45, -29, 2  49, -35, 6  55, -23, 2 | .023  .024  .024 |
| Thalamus | Right | 62 | 883.0  723.2 | 3 -17,-12  5, -25, -10 | .026  .046 |
| Brainstem |  | 48 | 818.3 | -7, -31, -8 | .032 |
| Putamen | Left | 23 | 730.8 | -33, -3, -8 | .045 |
| **Tremor amplitude-related activity (Tremor, averaged over blocks and sessions; ROI analysis)** | | | | | |
| **Anatomical location (area)** | **Hemisphere** | **Cluster size (voxels)** | **TFCE value** | **MNI coordinates peak voxel (x, y, z)** | ***p* (FWE)** |
| Motor cortex (4A) | Contralateral | 395 | 680.9  393.8  311.8 | -35, -29, 58  -44, -23, 56  38, -19, 52 | <.001  <.001  .001 |
| Cerebellum | Ipsilateral | 87 | 134.4  122.6 | 19, -49, -24  11, -51, -20 | .001  .002 |
| Thalamus (VLpv) | Contralateral | 21 | 24.4 | -19, -19, 8 | .070 |

The upper part of the table shows the activity related to general effects of the cognitive load task during fMRI, at the whole-brain level. The lower part of the table shows tremor amplitude-related activity across conditions and sessions, using regions of interest from previous studies and using small volume correction. For both analyses, TFCE was performed, and a threshold of p<.05 FWE-corrected at voxel level was used to determine significance. Anatomical labels were determined with the SPM Anatomy Toolbox^13^. TFCE=Threshold-free cluster enhancement, FWE=Family-wise error.

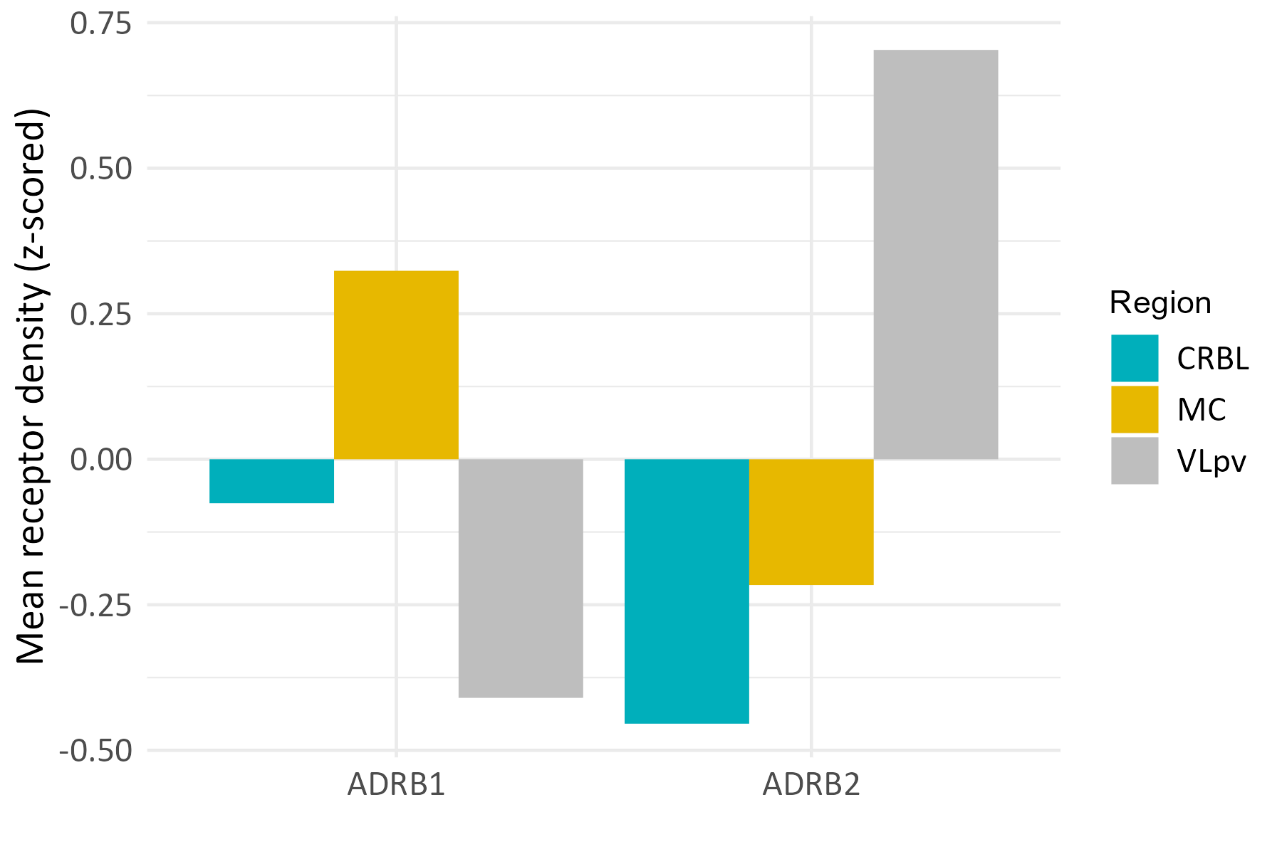

**Supplementary figure. Genetic expression of noradrenergic receptors in cerebral tremor network.** This figure shows the genetic expression of noradrenergic beta-1 and beta-2 receptors (ADRB1 and ADRB2; calculated from https://neurosynth.org/genes) in the cerebello-thalamo-cortical network. CRBL = cerebellum, MC = motor cortex, VLpv = ventrolateral nucleus of the thalamus, pars ventralis.
